# Validation of algorithms identifying diagnosed Obstructive Sleep Apnoea and narcolepsy in coded primary care and linked hospital activity data in England

**DOI:** 10.1101/2025.03.10.25323660

**Authors:** Helen Strongman, Sofia H. Eriksson, Kwabena Asare, Michelle A. Miller, Martina Sykorova, Hema Mistry, Kristin Veighey, Charlotte Warren-Gash, Krishnan Bhaskaran

## Abstract

**Purpose:** To assist sleep epidemiology research, we created and tested the accuracy of five algorithms identifying diagnosed Obstructive Sleep Apnoea (OSA) and narcolepsy in routinely collected data from England (01/01/1998-29/03/2021).

**Methods:** The primary algorithm identified the first coded record in Clinical Practice Research Datalink (CPRD) primary care or linked hospital admissions data as an incident diagnosis of OSA (n=92,222) or narcolepsy (n=1,072). Alternative algorithms required codes in CPRD, both datasets, or an additional proximate possible-sleep-related outpatient visit or excessive daytime sleepiness drug prescription (narcolepsy only). Staff in 73/1,574 CPRD practices completed online questionnaires for a convenience sample of 144 OSA and 101 narcolepsy cases. We estimated Positive Predictive Values (PPVs) describing the proportion of cases confirmed by a gold standard hospital specialist diagnosis, the percentage of gold standard cases from the primary algorithm retained with alternative algorithms, and time between specialist and recorded diagnosis dates.

**Results:** Using the primary algorithm, the PPV (95% CI) was 75.3% (69.2-81.3) and 65.2% (57.0-73.4) for OSA and narcolepsy, respectively: 80.6% and 62.7% of confirmed cases were recorded within 6 months of the specialist diagnosis. The CPRD-only algorithm increased the PPV to 85.3 (77.3-91.4, OSA) and 71.0 (58.8-81.3, narcolepsy) and retained high proportions of gold standard cases from the primary algorithm. Requiring additional outpatient or prescribing data increased PPVs substantially and improved diagnostic date accuracy for OSA but omitted a high proportion of gold standard cases.

**Conclusion:** Highly accurate OSA diagnoses can be identified in routinely collected data. Recorded cases of narcolepsy are moderately accurate, but diagnosis dates are not.

**Highlights:**

- Primary care records of OSA accurately represent hospital specialist diagnoses.
- Primary care records of narcolepsy are mostly accurate but diagnosis dates are not.
- More stringent definitions improve accuracy but identify fewer cases.
- Epidemiologists should use the best algorithm for their question and state limitations.

## Background

Sleep is a vital function to human health and daily living. Sleep can be disrupted by multiple environmental, lifestyle and medical factors including the primary sleep disorders Obstructive Sleep Apnoea (OSA) and narcolepsy(1), both of which are predominantly diagnosed in hospital-based specialist sleep centres using laboratory-based and ambulatory sleep studies(2).

Epidemiological research into sleep and other clinical fields can use routinely collected clinical and administrative data from healthcare systems. The early adoption of Electronic Health Records (EHR) to inform clinical care in UK general practices has supported the creation of large, longitudinal research databases linked to administrative hospital activity data(3). Research data are restricted to structured data fields including coded diagnoses from primary care and hospital admissions activity data. Coded diagnostic data are not routinely collected in hospital outpatient activity data. Studies investigating conditions treated in the hospital outpatient setting therefore rely on accurate coding of medical diagnoses by general practice staff based on receipt of clear information from hospital specialists, and the creation and use of high-quality code lists and algorithms by researchers to identify diagnoses(4). There is limited information assessing the quality of recording of OSA or narcolepsy diagnoses in routinely collected clinical data or the validity of codelists and algorithms that researchers use to identify these conditions.

To assess the utility of routinely collected data for sleep disorder research, we therefore investigated the validity of algorithms identifying OSA and narcolepsy diagnoses.

## Methods

### Study design

This validation study generated and tested the accuracy of five algorithms to identify diagnosed OSA and narcolepsy in de-identified primary care (CPRD Aurum) or linked hospital activity data (HES Admitted Patient Care – APC and outpatient) against a gold standard definition of diagnosis by a hospital specialist measured through a GP questionnaire study.

This study was approved by the London School of Hygiene & Tropical Medicine Ethics Committee (Ref 101296) and CPRD’s Research Data Governance process (protocol 22_001887). The study protocol is available online (5). CPRD supplies anonymised data for public health research; therefore, individual patient consent was not required for this study.

### Setting

UK General Practices provide a wide range of primary care services including the diagnosis, treatment and prevention of common conditions, and act as a gate keeper to specialist services. Practices collect data to support and audit these services in electronic health record software including EMIS Web. These data include Systematized Nomenclature of Medicine Clinical Terms (Snomed-CT) coded records of key clinical events and Dictionary of Medicines and Devices (DM+D) coded records of prescriptions. More detailed unstructured information describing clinical events are recorded in text boxes and by uploading documents such as letters received from hospitals and other care settings.

The Clinical Practice Research Datalink (CPRD) imports de-identified structured data from practices using EMIS software that have opted-in to providing data. Coded data is imported for all patients registered in these practices over time, except for people who have opted-out. Imported data include key demographic information such as sex and year of birth and a pseudonymised identifier that can only be decoded by GP practice staff. Personal identifiers, and unstructured data recorded in text boxes or uploaded documents are not imported. CPRD process imported data to form the CPRD Aurum database; this includes anonymised patient and practice identifiers(6).

Periodically, CPRD Aurum primary care data are linked to multiple datasets, including separate HES databases for each hospital setting (APC, Outpatient, Accident & Emergency), Office for National Statistics (ONS) death data, and area-based deprivation datasets through a trusted third party(7). Resultant linkage datasets include people with correct information necessary for data linkage at the time of processing.

We used the September 2023 CPRD Aurum build linked to the linkage dataset released in January 2022. The end of the data coverage period was 29/03/2021. The online questionnaire was administered through CPRD’s Providing Online Verification of Electronic Health Records service (CPRD PROVE Plus). Direct Object Identifiers (DOIs) for each dataset and links to information about CPRD PROVE are provided in Supplementary Appendix table 1.

### Participants

#### Study population

The study population included people registered in 1574 practices actively contributing to CPRD Aurum in May 2023 and included in the linkage dataset. To avoid duplication, practices that contributed to CPRD both before and after a practice merger were excluded. We further excluded people whose records failed CPRD’s data quality checks on recording and consistency of key variables(6), and for whom there was insufficient follow-up for an incident diagnosis to be identified (i.e. at least 90 days). The 90 day period was selected by visualising incidence rates of OSA and narcolepsy in the year following registration; records of OSA and narcolepsy during this time are likely to reflect earlier diagnoses (8) (Supplementary Appendix figure 1).

#### Incident sleep disorder cohorts

We used lookups provided by CPRD and NHS England to develop codelists including all codes for sleep apnoea and narcolepsy/cataplexy and recorded our decisions in a checklist(4) (Table 1). We developed and applied algorithms to identify incident OSA and narcolepsy cohorts and recorded diagnosis dates using these codelists. We first included people with a coded record of OSA or unspecified sleep apnoea or narcolepsy in CPRD Aurum or HES APC data. The date of the first inclusion record was assigned as the recorded diagnosis date. We then excluded people who were under 18 at diagnosis or had a prior record of central or primary sleep apnoea prior to or at diagnosis from the sleep apnoea cohort. To exclude prevalent diagnoses, we excluded people with a recorded diagnosis date before or within the first 90 days of practice registration from both cohorts.

**Table 1:**
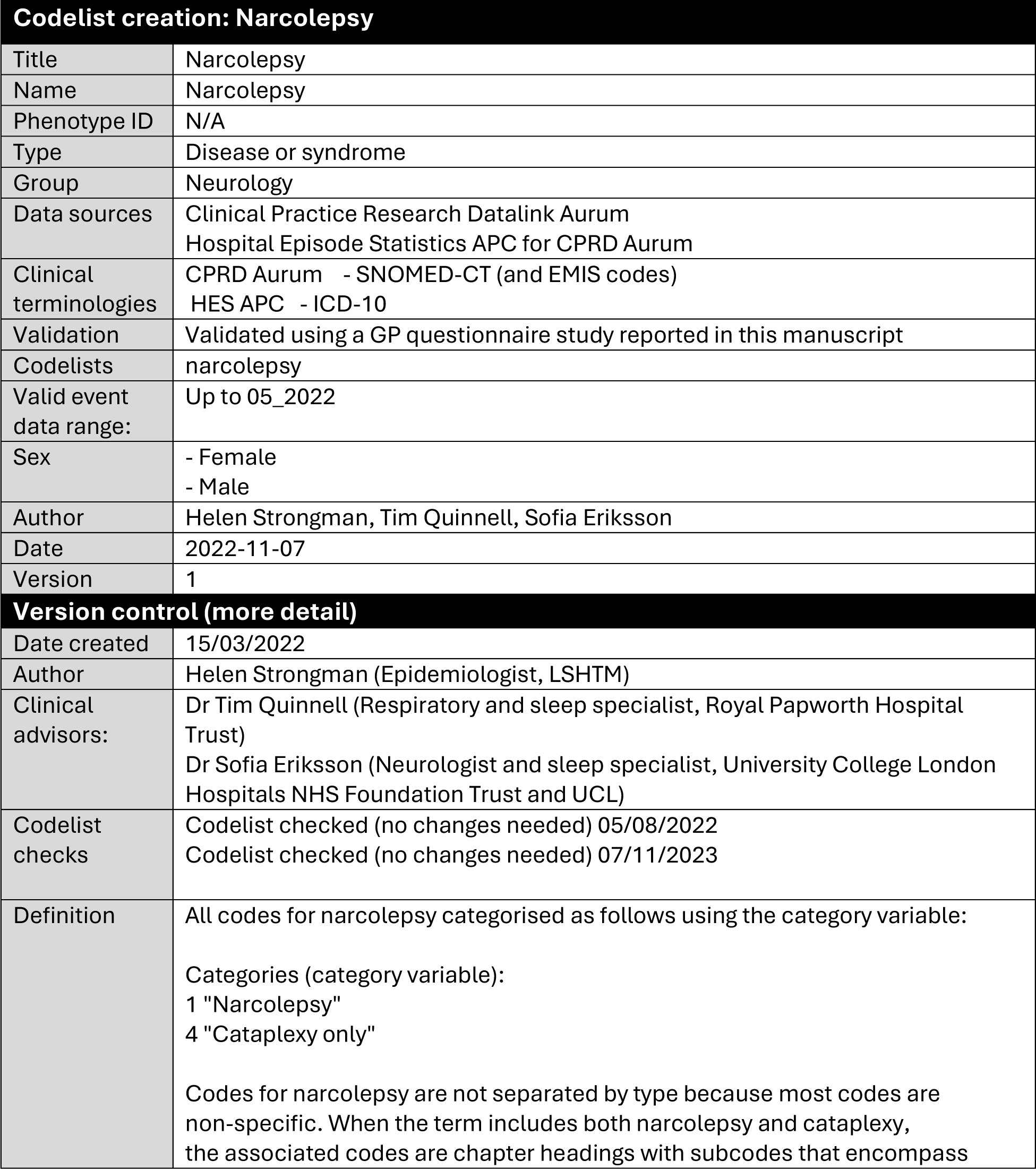
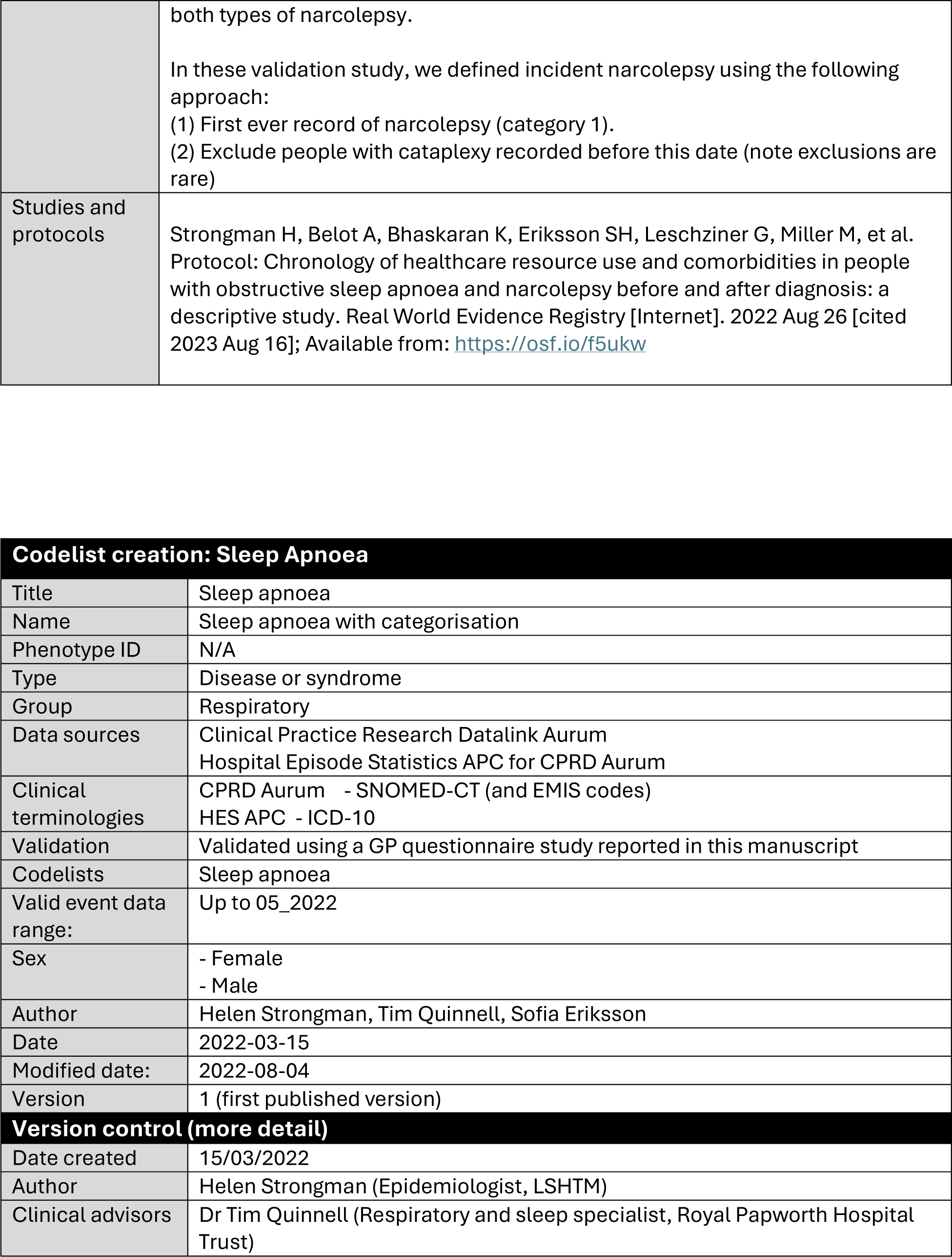
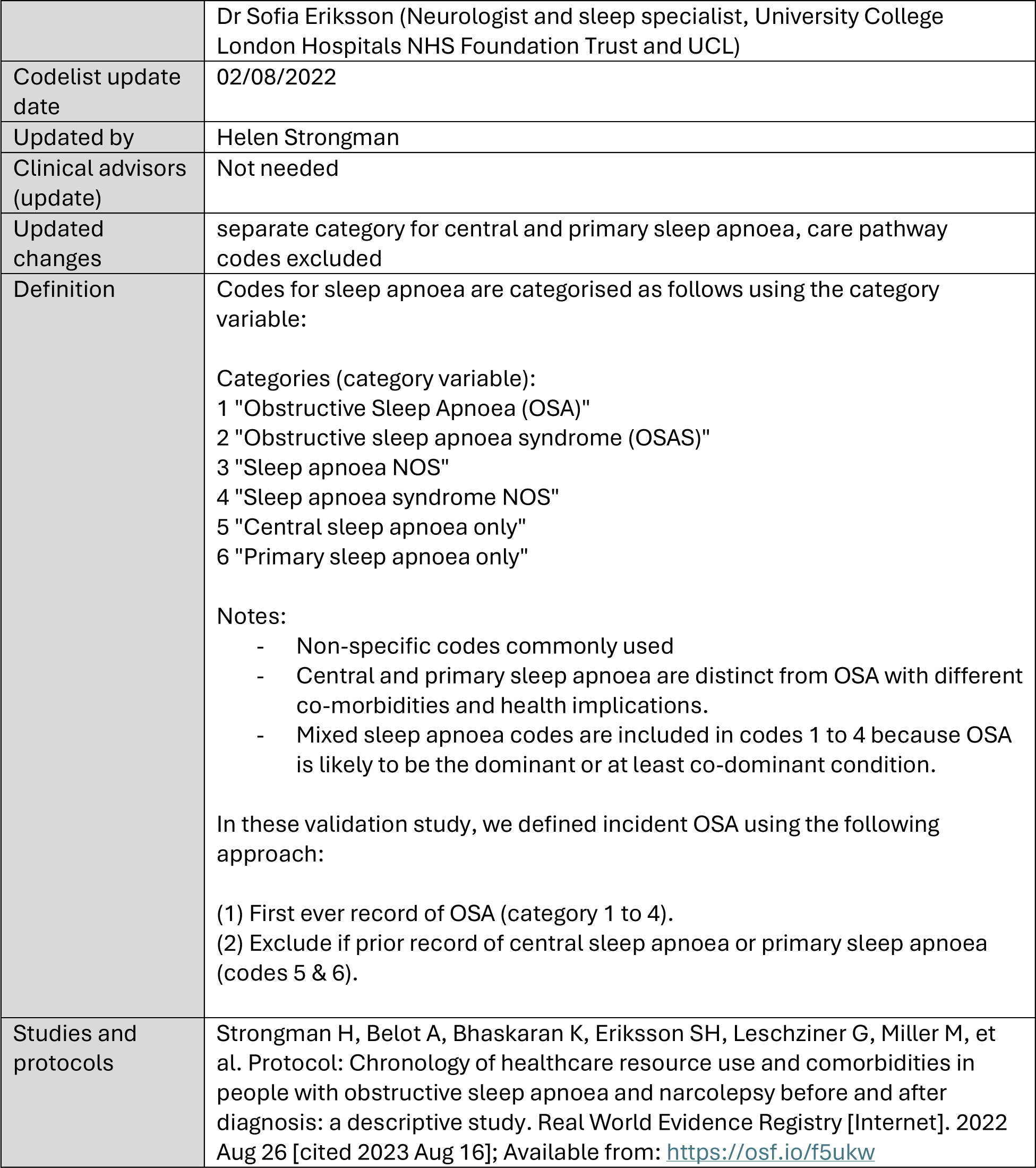
Checklists describing codelist creation methods for OSA and narcolepsy.

#### Sample size considerations

We used convenience sampling to collect data for a minimum of 100 cases in each cohort with each recruited practice completing three to four questionnaires. This approach balanced the scientific requirement to produce estimates with reasonable precision with practical considerations related to practice recruitment. The sample size calculation is included in Appendix text 1.

#### Recruitment sample

We restricted the incident cohort to a recruitment sample consisting of one or two narcolepsy cases and two OSA cases per practice. People registered in practices with no narcolepsy cases were excluded. Random sampling was used to select two cases of each sleep disorder in practices with additional cases.

We sent a list of CPRD Aurum patient identifiers, practice identifiers, and diagnosis dates to CPRD for each sleep disorder cohort. CPRD further eliminated practices that have recently stopped contributing data, those that have stated that they do not want to participate in research studies, and practices from a small number of Clinical Research Networks (CRNs) with no or more complex information governance processes in place for CPRD PROVE studies.

#### Validation sample

All remaining practices were then invited to sign up to a CPRD-web based agreement for this study on 18/03/2024 and for a member of practice staff to complete questionnaires for all cases in the recruitment sample for their practice on a first come, first served basis. Each practice received £110 per completed questionnaire, with an additional £110 incentive for completing all assigned questionnaires within one month.

The questionnaire was closed on 10/06/2024 when the minimum sample size of 100 completed questionnaires for the narcolepsy sample was reached. The resulting narcolepsy and OSA samples are referred to as the validation samples.

#### Validation questionnaire

The questionnaire was designed and reviewed in a text format by the full study team including epidemiologists, statisticians, sleep disorder clinicians and GPs. Following scientific approval of the published protocol, changes were made based on testing and narrative feedback by a General Practitioner (KV) and recommended adaptations for online questionnaire design by the CPRD PROVE team. The questionnaire was hosted on RedCap and tested by the study team and CPRD PROVE team. The system allowed respondents to indicate uncertainty in comments boxes instead of answering otherwise compulsory questions; missing values were returned for these responses and reset to “uncertain” for questions that included this response.

Figure 1 summarises the questionnaire structure. The full questionnaire is provided in Supplementary Appendix Text 2.

**Figure 1:**
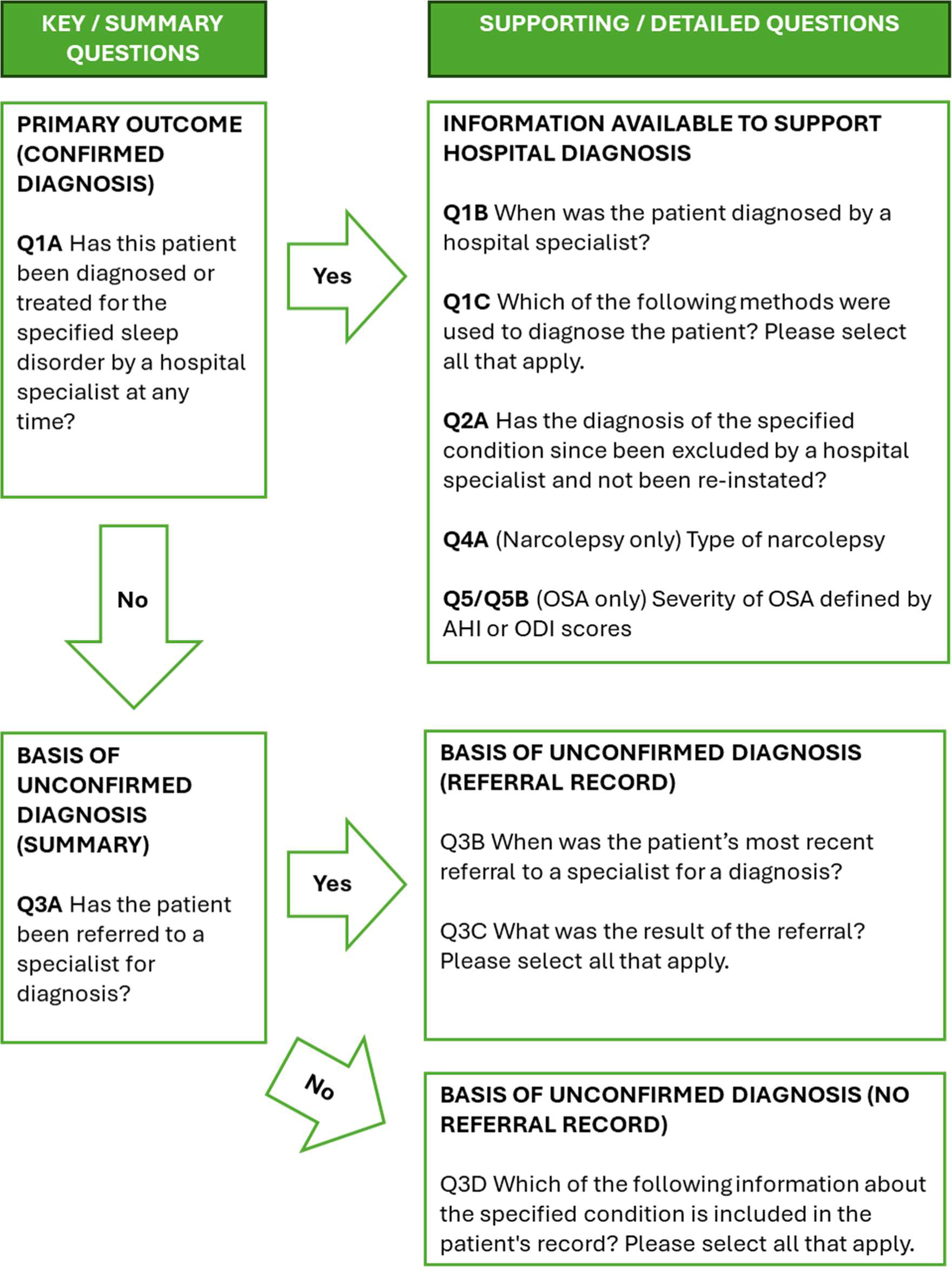
Validation questionnaire summary.

**Figure 2:**
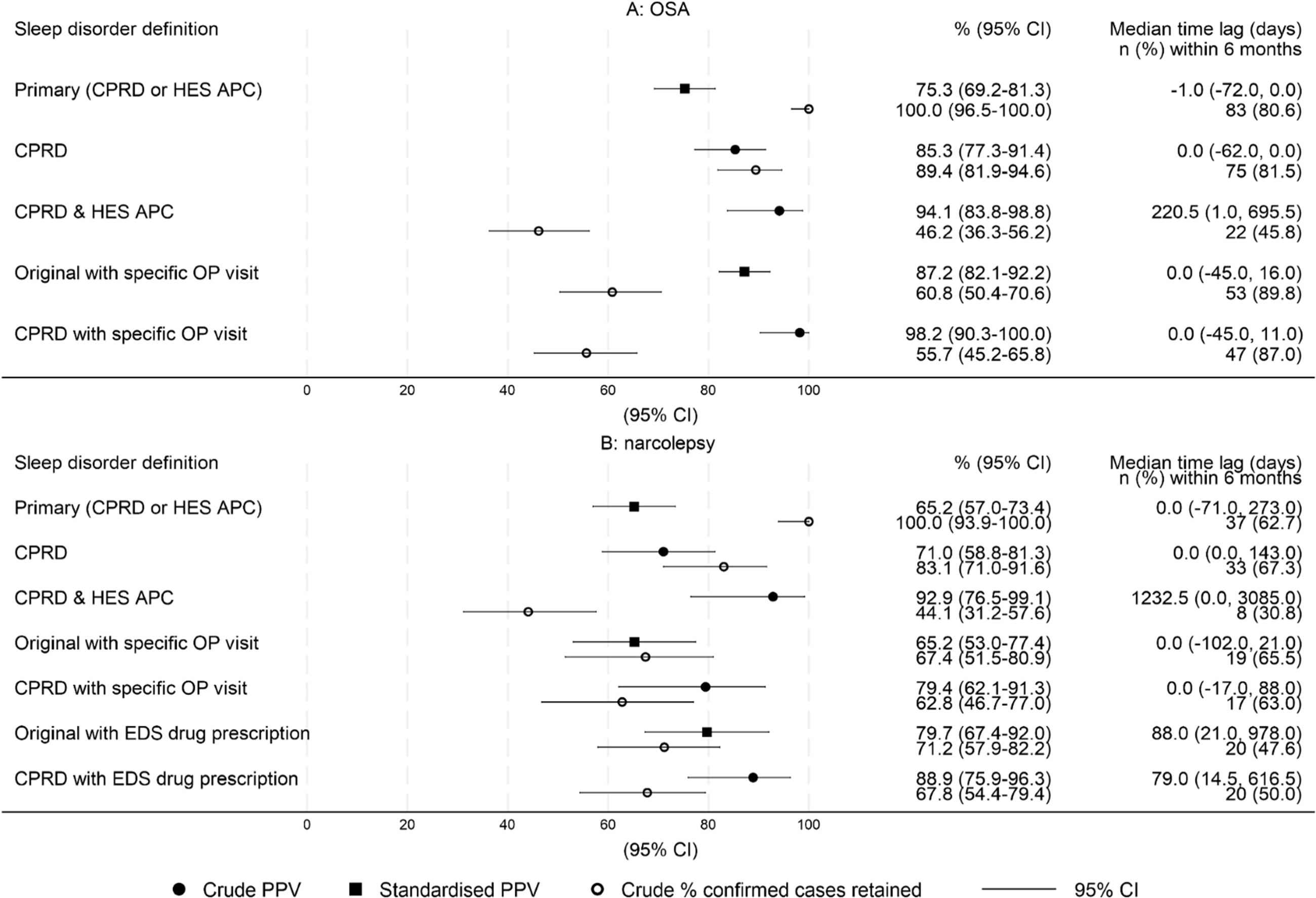
PPV, percentage of gold standard cases from the primary algorithm retained with stricter alternative algorithms, and time between recorded and specialist diagnosis dates, by algorithm. Caption: (1) Gold standard cases were identified as being diagnosed or treated by a hospital specialist in a GP questionnaire. (2) Substantial differences were observed in the proportion of cases identified in Clinical Practice Research Datalink (CPRD) or Hospital Episode Statistics Admitted Patient Care data (HES APC) in the incident cohort versus the validation sample and gold standard cases versus false positive cases. Standardised PPVs for the primary algorithm are therefore directly standardised to match the distribution of the incident cohort. (3) Gold standard cases retained are included in both the primary and alternative algorithm. (4) Median time lag, positive = recorded diagnosis date late.

### Variables

Full variable definitions are provided in Supplementary Appendix Table 2. All codelists are available at [add DOI before full publication]. Data management and statistical analyses were completed in Stata MP, version 17. Full programming code is available at https://github.com/hstrongman/OSA-narc_CPRD_validation/

#### Validation questionnaire variables

We designated gold standard diagnosed cases as those confirmed to be diagnosed or treated by a hospital specialist (Q1a “Has this patient been diagnosed or treated for the specified sleep disorder (see above) by a hospital specialist at any time?”=yes). The specialist diagnosis date was captured in Q1b “When was the patient diagnosed by a hospital specialist?” for gold standard cases. Further questions were designed to assess methods used to diagnose gold standard cases and the origin of false positive diagnoses. Data transformations for further descriptive variables are described in Supplementary Appendix Table 2.

#### Sleep disorder data and demographic variables

We defined the following variables describing data used to identify sleep disorder cases: category of diagnosis code (OSA only), origin of diagnosis code (CPRD and/or HES APC), calendar year at the recorded diagnosis and estimated person-years of follow-up prior to the recorded diagnosis. To support potential alternative sleep disorder algorithms, we additionally measured Excessive Daytime Sleepiness (EDS) drug prescriptions recorded at any time (narcolepsy only) and proximate HES outpatient visits (overall and sleep-related) in the 6 months before or after the recorded diagnosis. Sleep-related visits included those to outpatient clinics lead by neurology, respiratory, paediatrics, ENT and anaesthetics consultants. All analyses using HES outpatient data were restricted to people included in linkage processing for the HES outpatient data whose recorded diagnosis date was within the HES outpatient data coverage period (01/04/2003-30/10/2020). EDS drug prescriptions were identified in CPRD Aurum data and included prescriptions for modafinil, methylphenidate or dexamfetamine (i.e. drugs that are typically prescribed as 1^st^ or 2^nd^ line treatment).

Further demographic data defined using CPRD data included age at recorded diagnosis, sex, Body Mass Index (OSA only), ethnicity, practice area-based deprivation and urban-rural status, and practice size.

#### Alternative sleep disorder algorithms

We developed and applied alternative incident sleep disorder algorithms to the validation sample. These identified subsets of the cohort that met more stringent criteria and were hypothesised to reduce false positives, increasing the PPV, while reducing the number of gold standard cases identified. Pre-planned alternative algorithms included: coded records in CPRD data (no requirement for a code in HES APC data) and coded records in both CPRD and HES APC data. We explored possible alternative algorithms requiring both coded diagnoses and either EDS drug prescriptions (narcolepsy only) or proximate outpatient visits as defined above. Recorded diagnosis dates for all algorithms were set to the first date when all requirements were met.

## Statistical methods

### Descriptive and exploratory analyses

We compared sleep disorder data and demographic characteristics recorded in the incident cohorts and validation samples, and in gold standard and false positive cases within the validation study. Sleep disorder data and demographic variables whose distribution differed substantially in both comparisons were identified as standardisation variables. Possible alternative sleep disorder algorithms were confirmed for variables where distributions differed between gold standard and false positive cases.

### Validation estimates

We estimated the PPV for each sleep disorder algorithm by dividing the number of cases confirmed in the gold standard definition (i.e. those confirmed by GP staff to have been diagnosed/treated by a hospital specialist) by the total number of cases identified using the algorithm. PPVs for standardisation variables were directly standardised to match the distribution in the incident cohorts. We additionally estimated the percentage of gold standard cases identified using the primary algorithm that were retained when using the stricter alternative algorithms (gold standard cases meeting criteria for alternative algorithm/gold standard cases for primary algorithm). Exact binomial methods were used to estimate 95% confidence intervals.

To compare PPVs between demographic groups, we fitted generalized linear models with a binomial distribution and robust standard errors. This approach estimates relative differences in PPVs between groups.

Numbers representing 1 to 4 people, are reported as <5, in line with CPRD policy.

## Results

### Participants

From a study population of over 10 million people still registered in active CPRD Aurum practices at the last data collection date, 92,222 and 1,072 individuals were identified as being diagnosed with incident OSA or narcolepsy, respectively, while registered in the practice (Supplementary Appendix figure 2). The recruitment sample included 1,288 people with OSA and 901 with narcolepsy, for whom 144 and 101 questionnaires were completed by general practice staff in 73 practices.

### Descriptive and exploratory analyses

The median (IQR) age of the OSA and narcolepsy validation samples was 52.8 (43.8, 60.9) and 40.1 (IQR 23.0, 48.8), respectively. 29.2% (49) of the OSA and 52.5% (53) of the narcolepsy validation samples were female. The majority (>85%) of both validation samples were white and 67.9% of the OSA validation sample was obese (Table 2).

**Table 2:** Sleep disorder data and demographic characteristics recorded in the full incident cohorts and validation samples for OSA and narcolepsy.

|  | Incident OSA | Validation OSA | Incident narcolepsy | Validation narcolepsy |
| --- | --- | --- | --- | --- |
| **N+** | 92,054 | 168 | 971 | 101 |
| **RECORDING OF SLEEP DISORDER DIAGNOSIS** |  |  |  |  |
| **Source of diagnostic code\*** |  |  |  |  |
| Primary care | 66,200 (71.9) | 106 (63.1) | 655 (67.5) | 62 (61.4) |
| Inpatient hospital activity data | 25,854 (28.1) | 62 (36.9) | 316 (32.5) | 39 (38.6) |
| **Most specific sleep apnoea code type recorded at index** |  |  |  |  |
| OSA code | 55,447 (60.2) | 110 (65.5) |  |  |
| sleep apnoea | 36,607 (39.8) | 58 (34.5) |  |  |
| **Person-years before diagnosis/index** |  |  |  |  |
| Mean (SD) | 16.6 (13.4) | 16.8 (13.7) | 12.9 (11.4) | 13.8 (11.5) |
| Median (IQR) | 13.7 (5.7, 24.1) | 14.9 (5.7, 23.4) | 9.6 (4.3, 18.3) | 12.2 (4.9, 19.3) |
| **Calendar year at index** |  |  |  |  |
| Mean (SD) | 2014.4 (5.1) | 2014.2 (5.1) | 2012.7 (6.0) | 2012.9 (6.6) |
| Median (IQR) | 2016 (2011, 2018) | 2015.5 (2011, 2018) | 2014 (2008, 2018) | 2014 (2010, 2019) |
| **ADDITIONAL SUPPORTING INFORMATION IN ROUTINELY COLLECTED DATA** |  |  |  |  |
| **Outpatient visit within 6 months of diagnosis (HES OP data)** |  |  |  |  |
| Linked OP data available | 79,361 (86.2) | 150 (89.3) | 800 (82.4) | 75 (74.3) |
| All | 72,156 (90.9) | 129 (86.0) | 707 (88.4) | 64 (85.3) |
| Neurology | 4,796 (6.0) | 9 (6.0) | 328 (41.0) | 35 (46.7) |
| Respiratory | 39,902 (50.3) | 65 (43.3) | 241 (30.1) | 23 (30.7) |
| Paediatric (NOS) | 63 (0.1) | 0 (0.0) | 103 (12.9) | 8 (10.7) |
| Ear Nose & Throat | 15,226 (19.2) | 25 (16.7) | 71 (8.9) | <5 (<6.7) |
| Anaesthetics | 63 (0.1) | 0 (0.0) | 103 (12.9) | 8 (10.7) |
| Sleep-related consultants combined | 50,143 (63.2) | 83 (55.3) | 562 (70.2) | 50 (66.7) |
| **Excessive Daytime Sleepiness drug prescriptions** |  |  |  |  |
| EDS drug prescription ever |  |  | 545 (56.1) | 49 (48.5) |
| **Days between recorded diagnosis date & 1st EDS drug prescription (positive = recorded first)** |  |  |  |  |
| Mean (SD) |  |  | -85.0 (1221.2) | -222.7 (819.2) |
| Median (IQR) |  |  | 37.0 (0.0, 203.0) | 16.0 (-165.0, 158.0) |
| **Recorded diagnosis compared to date of 1st EDS drug prescription** |  |  |  |  |
| > 6 months before |  |  | 90 (9.3) | 11 (10.9) |
| within 6 months |  |  | 308 (31.7) | 27 (26.7) |
| > 6 months after |  |  | 146 (15.0) | 11 (10.9) |
| missing |  |  | 427 (44.0) | 52 (51.5) |
| **CHARACTERISTICS** |  |  |  |  |
| **Age at diagnosis (years)** |  |  |  |  |
| Mean (SD) | 52.4 (12.5) | 51.5 (12.6) | 37.5 (18.5) | 37.6 (17.2) |
| Median (IQR) | 52.4 (43.8, 61.0) | 52.8 (43.8, 60.9) | 36.9 (23.8, 51.2) | 40.1 (23.0, 48.8) |
| **Sex** |  |  |  |  |
| male | 62,603 (68.0) | 119 (70.8) | 464 (47.8) | 48 (47.5) |
| female | 29,451 (32.0) | 49 (29.2) | 507 (52.2) | 53 (52.5) |
| **Body Mass Index** |  |  |  |  |
| Under/normal weight | 8,045 (8.7) | 17 (10.1) |  |  |
| Overweight | 20,935 (22.7) | 32 (19.0) |  |  |
| Obesity class I | 22,840 (24.8) | 49 (29.2) |  |  |
| Obesity class II | 16,669 (18.1) | 30 (17.9) |  |  |
| Obesity class III+ | 19,234 (20.9) | 35 (20.8) |  |  |
| missing | 4,331 (4.7) | 5 (3.0) |  |  |
| **Ethnicity** |  |  |  |  |
| 0. White | 80,199 (87.1) | 146 (86.9) | 814 (83.8) | 88 (87.1) |
| 1. South Asian | 5,682 (6.2) | 9 (5.4) | 36 (3.7) | <5 (<5.0) |
| 2. Black | 3,456 (3.8) | 11 (6.5) | 77 (7.9) | 6 (5.9) |
| 3. Other | 1,385 (1.5) | <5 (<3.0) | 14 (1.4) | <5 (<5.0) |
| 4. Mixed | 967 (1.1) | <5 (<3.0) | 21 (2.2) | 0 (0.0) |
| missing | 365 (0.4) | 0 (0.0) | 9 (0.9) | <5 (<5.0) |
| **Carstairs quintile** |  |  |  |  |
| 1 (least deprived) | 11,129 (12.1) | 38 (22.6) | 95 (9.8) | 24 (23.8) |
| 2 | 18,119 (19.7) | 31 (18.5) | 171 (17.6) | 18 (17.8) |
| 3 | 20,724 (22.5) | 39 (23.2) | 234 (24.1) | 26 (25.7) |
| 4 | 20,450 (22.2) | 30 (17.9) | 215 (22.1) | 18 (17.8) |
| 5 (most deprived) | 21,632 (23.5) | 30 (17.9) | 256 (26.4) | 15 (14.9) |
| **Urban Rural** |  |  |  |  |
| urban | 77,508 (84.2) | 131 (78.0) | 846 (87.1) | 78 (77.2) |
| rural | 14,282 (15.5) | 37 (22.0) | 120 (12.4) | 23 (22.8) |
| missing | 264 (0.3) | 0 (0.0) | 5 (0.5) | 0 (0.0) |
| **Practice size** |  |  |  |  |
| Mean (SD) | 42006.6 (34157.0) | 40362.9 (32850.8) | 41839.8 (33948.9) | 43438.7 (37083.2) |
| Median (IQR) | 32319 (22644, 47644) | 30927 (20682, 44124) | 32433 (22517, 46802) | 31759 (20749, 48163) |

\*A small number of people had a code in both sources on the index date (<2%). These are coded as CPRD Aurum to avoid small cell counts

Sleep disorder diagnoses were recorded between 1998 and 2021 with most codes derived from the CPRD data; non-specific sleep apnoea codes were used for one third or the OSA cohort/sample. The majority of people in the validation sample were eligible for linkage to HES outpatient data. Of these, 86.0% (129, OSA) and 85.3% (64, narcolepsy) had a proximate outpatient visit and 55.3% (OSA) and 66.7% (narcolepsy) had proximate possible sleep-related outpatient visit. 48.5% (n=49) of the validation sample had at least one prescription of an EDS drug in their primary care record (Table 2).

Diagnoses recorded in HES APC were more prevalent in the validation samples (36.9% OSA, 38.6% narcolepsy) than the incident cohorts (28.1% OSA, 32.5% narcolepsy) (table 2) and less prevalent in gold standard cases (23.1% OSA, 28.8% narcolepsy) than false positive cases (62.5% OSA, 52.4% narcolepsy) (Supplementary Appendix Table 3). The source of the diagnosis code was therefore identified as a stratification variable.

Alternative algorithms including proximate outpatient visits to a possible sleep-related specialist and an EDS prescription were selected as substantial differences in the distribution of these variables were observed in gold standard vs false positive cases in the OSA and narcolepsy validations samples, respectively (Supplementary Appendix Table 3).

### Outcomes and estimation

For each algorithm, Figure 3 describes the PPV, percentage of gold standard cases from the primary algorithm that were retained using the stricter alternative algorithms, and time difference between the recorded and specialist diagnosis date. Using the primary algorithm, the standardised percentage (95% CI) of gold standard cases was 75.3 (69.2-81.3) for OSA and 65.2 (57.0-73.4) for narcolepsy. The median number of days between the recorded and specialist diagnosis date, and percentage of gold standard cases recorded within 6 months of the specialist diagnosis, was -1.0 (IQR -72.0,0.0, 80.6%) for OSA and 0.0 (IQR -71.0, 273.0, 62.7%) for narcolepsy. Using the CPRD only algorithm increased the percentage (95% CI) of gold standard cases to 85.3 (77.3-91.4) for OSA and 71.0 (58.8-81.3) for narcolepsy, retained 89.4% (95% CI 81.9-94.6) of gold standard OSA and 83.1% (95% CI 71.0-91.6) of gold standard narcolepsy cases from the primary algorithm, and made little difference to the accuracy of the diagnosis date.

For OSA, requiring CPRD data with a proximate possible sleep-related outpatient visit to identify diagnosed cases provided the best balance between maximising the PPV (98.2%, 95% CI 90.3-100) while retaining the highest proportion of gold standard cases from the primary algorithm OSA 55.7% (95% CI 45.2-65.8). Using this algorithm also increased the percentage of recorded cases within 6 months of the specialist diagnosis to 86.9%. For narcolepsy, requiring CPRD data with an EDS drug prescription provided the best balance; PPV 88.9% (95% CI 75.9-96.3), gold standard cases retained 67.8% (95% CI 54.4-79.4) with a further reduction in the accuracy of the diagnosis date.

There is little evidence that the PPV differed between patient characteristics, except for Body Mass Index for OSA: PPV (95% CI) 67.1% (55.8-77.1) for the obesity class I and lower category and 82.8% (70.6-91.4) for the obesity class II and above category. There is weak evidence that the crude PPV for narcolepsy was lower in later calendar-years. (Supplementary Appendix table 4).

### Descriptive analyses of questionnaire responses

Table 3 describes responses to the full validation questionnaire and sample and Supplementary table 5 summarises key information across all algorithms.

**Table 3:**
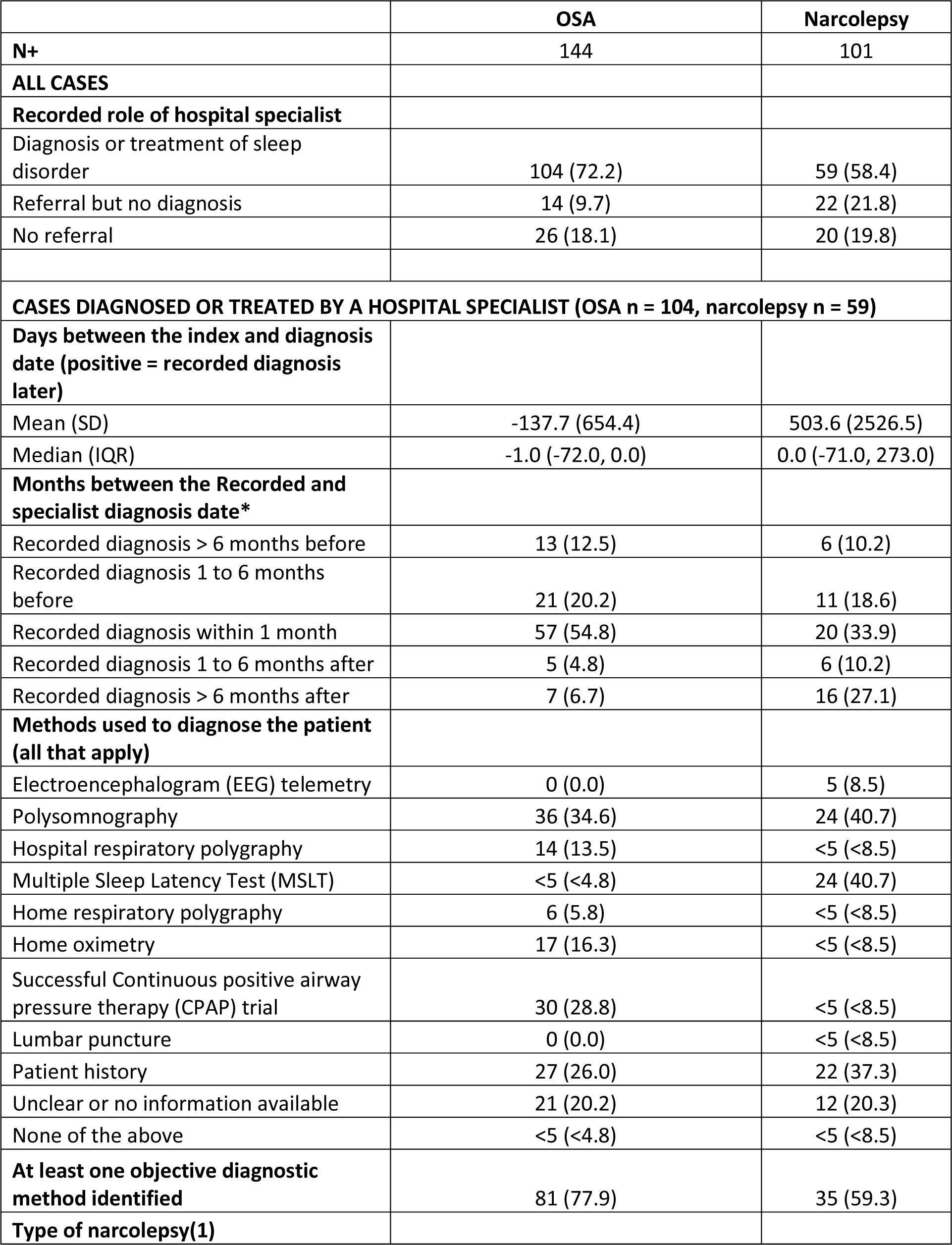
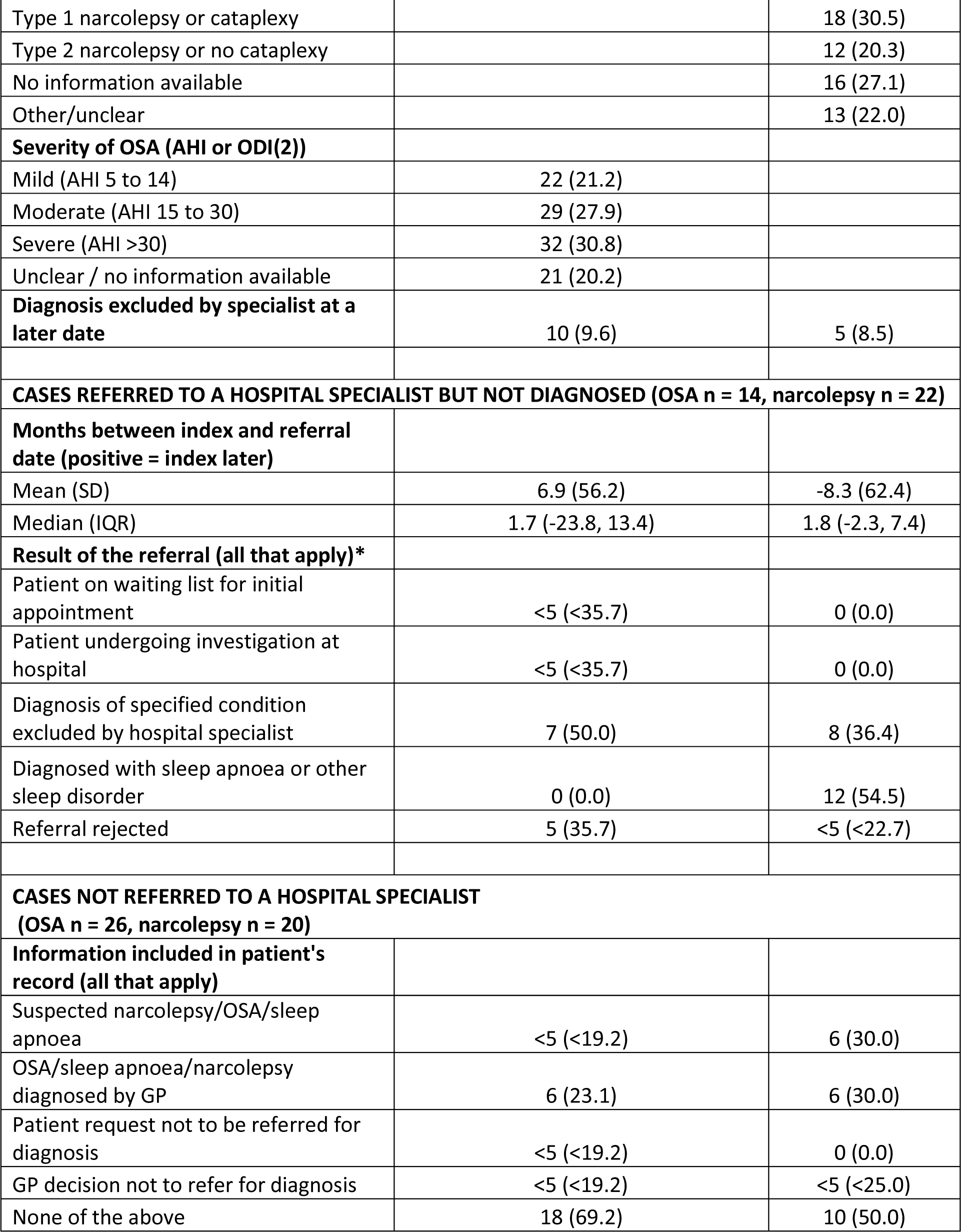
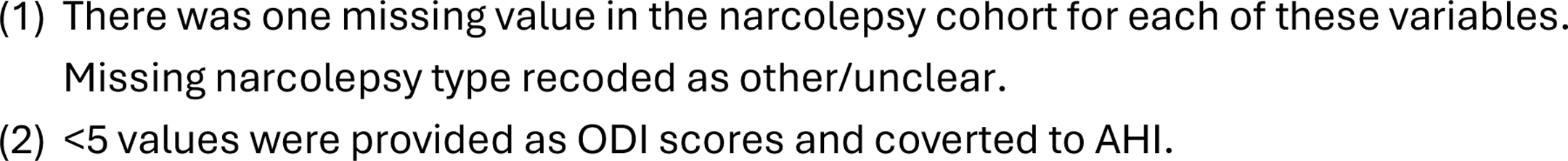
Validation questionnaire responses.

At least one objective diagnostic method was identified for 77.9% and 59.3% of gold standard OSA and narcolepsy cases, respectively. This proportion was similar across all algorithms for OSA but increased with more stringent narcolepsy algorithms (max 70.4% CPRD with outpatient visit to sleep-related specialty). The type of narcolepsy was identified in half of cases (30.5% type 1, 20.3% type 2). OSA severity was recorded for nearly 80% of cases with a fairly even split between mild, moderate and severe cases. The original hospital specialist diagnosis was excluded at a later date for 9.6% (n=10) of OSA and 8.5% (n=5) of narcolepsy gold standard cases reflecting the complexity in diagnosing these conditions and potential alleviation of OSA symptoms.

9.7% (14) and 21.8% (22) of people in the OSA and narcolepsy validation samples, respectively, were referred to hospital specialists but not diagnosed. Amongst these cases, the diagnosis had been excluded or another sleep disorder had been diagnosed in 50% of OSA and nearly all narcolepsy cases.

There was no evidence of referral to a hospital specialist for 18.1% (26) and 19.8% (20) of people in the OSA and narcolepsy validation samples, respectively. The reason for this is unclear in 69.2% (OSA) and 50% (narcolepsy) of cases. Others were either suspected cases, diagnosed by a GP, or not referred for diagnosis.

## Discussion

### Summary

We tested the accuracy of 5 algorithms to identify diagnosed OSA and narcolepsy in de-identified primary care (CPRD Aurum) and hospital activity data (HES APC and HES outpatient) against a gold standard definition of diagnosis by a hospital specialist. The primary algorithm of a coded record in either CPRD Aurum or HES APC data had the lowest PPV for both OSA (75.3 95% CI 69.2-81.3) and narcolepsy (65.2 95% CI 57.0-73.4). The accuracy of the diagnostic date was moderate for OSA and poor for narcolepsy. Using CPRD data only increased the PPV to 85.3 (95% CI 77.3-91.4) for OSA and 71.0 (95% CI 58.8-81.3) for narcolepsy, while losing approximately 10 to 20% of gold standard cases from the primary algorithm. More stringent algorithms that achieved the best balance between increasing the PPV while retaining gold standard cases required CPRD data with a proximate possible sleep-related outpatient visit for OSA (PPV 98.2%, 95% CI 90.3-100.0) and requiring CPRD data with a prescription record for an EDS drug for narcolepsy (PPV 88.9% 95% CI 75.9-96.3).

### Strengths and limitations

The key strength of our study is that we were able to assess whether coded incident diagnoses of OSA and narcolepsy in de-identified data represented specialist diagnoses using additional information that is available in the full primary care record, through a structured questionnaire. Setting hospital specialist diagnoses as a gold standard matches diagnosis and care pathways for both OSA and narcolepsy, both of which require specialist care. To enable our algorithms to be used in future studies, we have recorded methods used to construct code lists for each source(4), and our analysis code.

As we did not validate records with no recorded diagnosis, we were unable to estimate negative predictive values (i.e. the proportion of people without a recorded diagnosis who have not been diagnosed) or the sensitivity of our algorithms (i.e. the proportion of gold standard (hospital specialist) diagnosed OSA and narcolepsy cases in the study population that we identified with our algorithms. The negative predictive value of our algorithms is likely to be high as the ratio of diagnosed to undiagnosed people in the population is low, even in high risk OSA groups. Our analysis plan considered uncertainty and potential selection bias introduced through our convenience sampling strategy. This strategy allowed us to reach our target sample within 3 months, include a large number of practices, and provide reasonably certain estimates and detailed questionnaire responses for the primary algorithm. Wider confidence intervals for narrower alternative algorithms mean that differences between algorithms may be due to chance alone, and small cell count requirements prevent us from describing responses in full. We identified higher proportions of cases recorded in HES APC data only in the validation sample, compared to the full eligible incident sample, as a potential selection bias. We mitigated this bias by stratifying PPVs by source for algorithms affected by this bias. We were unable to identify mechanisms of selection bias caused by unmeasured differences between the full incident sample and validation sample or by restricting our study to active practices and cases. Additionally, recording practices may have changed during the Covid-19 pandemic(9); this may explain our observation of weak evidence of a lower PPV for narcolepsy later in the study period.

Our validation method relies on specialists transferring accurate information to GPs and GP staff being able to find and interpret this information. Whilst the origin of the majority of false positive cases identified using the primary algorithm was confirmed in the questionnaire (e.g. diagnosis excluded after a referral), 45% (40) and 22.7% (18) of false positive OSA and narcolepsy cases, respectively, had no record of referral to a specialist and no clear reason for the condition being recorded. Algorithms using CPRD data only resulted in a higher PPV and lower proportion of false positive OSA cases with no referral to hospital specialists. This may reflect information not being transferred from hospitals to GPs, or GP staff completing this questionnaire not being able to find this information in scanned letters, resulting in false negative cases. This pattern persists for algorithms requiring outpatient visits and EDS drug prescribing by the general practice (narcolepsy only). Similarly, moderate proportions of objective testing for gold standard cases identified across narrow and broad algorithms suggests a lack of clear and detailed information being transferred to GPs from hospital specialists.

Furthermore, we assessed whether coded records accurately identified specialist diagnoses at the time of recording. Questionnaire responses indicated that 9.6% of OSA and 8.5% of narcolepsy diagnoses were excluded by a hospital specialist at a later date. These may be borderline cases for which symptoms vary over time or cases where OSA symptoms have been alleviated by lifestyle changes; diagnoses may also be excluded as knowledge increases or people are referred to tertiary sleep centres.

### Strengths and limitations in comparison to existing studies

There is minimal published evidence assessing the validity of OSA and narcolepsy records in routinely collected data. A validation study of recording of sleep disorders diagnosed in a single Canadian sleep centre found that non-specific coding of sleep disorders was common in all data sources, and particularly poor in inpatient data(10). It is possible that GPs and hospitals use similarly generic terms to record OSA and narcolepsy in England; these cases would be not be identified using our algorithms. In contrast, recording of sleep apnoea was found to be highly accurate in electronic health record data from US hospitals that participated in a sleep apnoea genetics study(11). The latter study included coded data from outpatient appointments. We found that requiring a proximate visit to one of four possible sleep related specialties improved the PPV for both sleep disorders, while substantially reducing the number of cases identified. Whereas US hospitals may be motivated to accurately record these data for re-imbursement purposes, recording of diagnostic data in English HES outpatient data is not mandated by NHS England and therefore highly incomplete. We were therefore unable to use this source to identify cases or validate cases identified in other sources.

A systematic review estimated a median PPV of 89% (95% CI 24-100%) for 183 different diagnoses in CPRD primary care data (linked data were not available at the time)(12). There was no clear pattern by ICD-10 Chapter and data for individual diagnoses or papers was not presented. Our estimated PPV for OSA using CPRD data only of 85.3% (95% CI 77.3%-91.4%) is close to the median whereas our estimated PPV for narcolepsy of 71.0% (95% CI 58.8-81.3%) is lower than median but well within the interquartile range.

A recent concordance study including cancer registration data in the gold standard algorithm estimated PPVs greater than 80% for CPRD records for each of the 20 most common cancers(13). PPVs were highest for common, clearly defined and well-understood cancers with higher survival rates; lack of familiarity with narcolepsy among GPs and the complexity of diagnosing the condition may therefore explain lower than average PPVs. In contrast to our findings, PPVs for cancer algorithms that included records in CPRD, HES or ONS mortality data were similar to CPRD only algorithms. Unlike our study, this concordance study did not rely on data transfer from hospitals to GP practices to measure gold standard cases. Our hypothesis that this biased the PPV downwards is further supported by a study by Winstone et al.(14). This study reported high accuracy albeit low completeness of childhood narcolepsy cases recorded in English HES data based on a review of clinical notes and investigation records from specialist centres by three narcolepsy experts. Alternatively, incorrect HES records of narcolepsy diagnoses may be less common in children.

### Meaning of the study and future research

Incident diagnoses recorded in primary care data in England are highly accurate for OSA and reasonably accurate for narcolepsy. However, diagnosis dates are moderately accurate for OSA and poorly recorded for narcolepsy. Our findings suggest that including coded records from hospital admissions data reduces the accuracy of the recorded diagnosis; however this may be due to limitations associated with information transfer between hospitals and primary care. Requiring a proximate possible sleep related outpatient visit or an EDS drug prescription for narcolepsy increases the accuracy of recorded diagnoses substantially but removes a high proportion of gold standard cases. As UK primary care EHR systems are designed to be inter-operable these findings are likely to generalise to other EHR systems (e.g. SystmOne) and nations of the UK.

Use of routinely collected data is often either the only way to study population based epidemiological questions, or a useful supplement to studies using more accurate prospectively collected data from smaller less representative samples of the population. When using routinely collected data from England’s NHS to study OSA or narcolepsy, epidemiologists should select the most appropriate algorithm for their study, include sensitivity analyses using alternative algorithms, and acknowledge limitations associated with measurement bias.

The recently released Sudlow report provides recommendations for improving the UK health data landscape(3). These recommendations include mandating the inclusion of diagnosis and procedural codes in national hospital outpatient episodes data, and automated coding of unstructured information from electronic health records. We believe that both of these recommendations would substantially improve case completeness and our ability to accurately identify OSA and narcolepsy cases. We further recommend that specialist sleep centres agree on a common use of ICD-10 coding in HES outpatient data to record suspected and diagnosed cases; this would support future epidemiological research. Audits of information transferred from hospital specialists to primary care(15), and improving medical education about sleep disorders(16), may be used to improve recording in primary care and future automated coding of unstructured data. More accurate recording of sleep disorders in general practice would directly improve the quality and safety of care in addition to supporting impactful research(17).

## Conclusion

Recorded diagnoses of two well-defined sleep disorders, OSA and narcolepsy, in routinely collected GP data are highly and reasonably accurate, respectively. Diagnosis dates are moderately accurate for OSA but poorly recorded for narcolepsy. Stricter algorithms using HES outpatient data and CPRD prescribing data (narcolepsy only) improve accuracy substantially, while missing high numbers of gold standard diagnosed cases.

Epidemiologists should use the algorithm most suited to their study and include sensitivity analyses using alternative algorithms. We support recommendations to mandate recording of diagnosis codes in hospital outpatient episodes data; this is likely to improve case ascertainment and accuracy.

## Data Availability

Data were obtained from the Clinical Practice Research Datalink (CPRD). CPRD is a research service that provides primary care and linked data for public health research. CPRD data governance, and the authors' own licence to use CPRD data, do not allow the authors to distribute or make available patient data directly to other parties. Researchers may apply for data access and must have their study protocol approved via CPRD's Research Data Governance Process.

## Acknowledgements

This study is based in part on data from the Clinical Practice Research Datalink obtained under licence from the UK Medicines and Healthcare products Regulatory Agency. Hospital Episodes Statistics Data and Office of National Statistics Mortality data were reused with the permission of The Health & Social Care Information Centre (All rights reserved, Copyright (2021)). The data are provided by patients and collected by the NHS as part of their care and support. We would like to thank Harley Kennedy and Iryna Brooks for managing data collection at CPRD. The interpretation and conclusions based in this study are those of the authors alone.

