## Supplementary Appendix for "Validation of algorithms identifying diagnosed Obstructive Sleep Apnoea and narcolepsy in coded primary care and linked hospital activity data in England"

### SA Text 1 : Sample calculation

A systematic review estimated a mean Positive positive predicted Value (PPV) of 90% for 357 studies validating code lists and algorithms used to identify medical conditions in CPRD data (Herrett et al). With a random sample size of 100 for each sleep disorder, the 95% confidence intervals (based on a normal distribution) would be 82.4% to 95.1%. Subset analyses would also have reasonable precision (e.g. expected 95% CI of 78.2% to 96.7% for a subset including 50 people).

Herrett E, Thomas SL, Schoonen WM, Smeeth L, Hall AJ. Validation and validity of diagnoses in the General Practice Research Database: a systematic review. British journal of clinical pharmacology. 2010 Jan;69(1):4–14.

### SA Text 2: Validation questionnaire

| **VALIDATION OF NARCOLEPSY AND OBSTRUCTIVE SLEEP APNOEA RECORDS** |
|---|
| The London School of Hygiene and Tropical Medicine is conducting a study to investigate the strengths and limitations of using Clinical Practice Research Datalink (CPRD) to study the epidemiology of narcolepsy and sleep apnoea. We have identified narcolepsy and Obstructive Sleep Apnoea (OSA) diagnoses in the database and linked hospital data (where available) by selecting the first coded record of either narcolepsy or OSA. We would like to collect data about information that supports these diagnoses so that we can understand how valid this method is. We would be very grateful if you could answer the following questions for the specified condition only. |
| *Please answer questions based on the following diagnosis:*<br>☐ **Narcolepsy**<br>☐ **Obstructive Sleep Apnoea (OSA)**<br>This may also be referred to as obstructive sleep apnoea syndrome (OSAS), or obstructive sleep apnoea hypopnoea syndrome (OSAHS). Non-specific codes for sleep apnoea may also be used in the primary care record with the type of sleep apnoea specified in the hospital letter. People with mixed or complex sleep apnoea have OSA whereas primary and central sleep apnoea are separate conditions. |

| NARCOLEPSY SURVEY FORM | |
|---|---|
| **Q1A.** | Has this patient been diagnosed or treated for the specified sleep disorder (see above) by a hospital specialist at any time?<br><br>☐ Yes *(Go to Q1B, Q1C and Q2A)*<br>☐ No *(Go to Q3A)* |
| **Q1B.** | When was the patient diagnosed by a hospital specialist?<br><br>☐ *(insert date DD/MM/YYYY – cannot choose a date in the future, the latest date should be current date)* |
| **Q1C.** | Which of the following methods were used to diagnose the patient? *(Please select all that apply)*<br><br>☐ Electroencephalogram (EEG) telemetry<br>☐ Polysomnography<br>☐ Hospital respiratory polygraphy<br>☐ Multiple Sleep Latency Test (MSLT)<br>☐ Home respiratory polygraphy<br>☐ Home oximetry<br>☐ Successful Continuous positive airway pressure therapy (CPAP) trial<br>☐ Lumbar puncture<br>☐ Patient history<br>☐ Unclear or no information available<br>☐ None of the above |
| **Q2A.** | Has the diagnosis of the specified condition since been excluded by a hospital specialist and not been re-instated?<br><br>☐ Yes *(Go to Q4)*<br>☐ No *(Go to Q4)* |
| **Q3A.** | Has the patient been referred to a specialist for diagnosis?<br><br>☐ Yes *(Go to Q3B and Q3C.)*<br>☐ No *(Go to Q3D.)* |
| **Q3B.** | When was the patient's most recent referral to a specialist for diagnosis?<br><br>☐ *(insert date DD/MM/YYYY – cannot choose a date in the future, the latest date should be current date)* |
| **Q3C.** | What was the result of the referral? *(Please select all that apply)*<br><br>☐ Patient on waiting list for initial appointment<br>☐ Patient undergoing investigation at hospital |

| | |
|---|---|
| | ☐ Diagnosis of specified condition excluded by hospital specialist<br>☐ Diagnosed with OSA (relevant when the specified sleep disorder is narcolepsy)<br>☐ Diagnosed with unspecified sleep apnoea<br>☐ Diagnosed with other sleep apnoea (Primary or central)<br>☐ Diagnosed with other sleep disorder<br>☐ Referral rejected |
| **Q3D.** | Which of the following information about the specified condition is included in the patient's record? *(Please select all that apply)*<br><br>☐ Suspected narcolepsy/OSA/sleep apnoea<br>☐ OSA/sleep apnoea/narcolepsy diagnosed by GP<br>☐ Patient request not to be referred for diagnosis<br>☐ GP decision not to refer for diagnosis<br>☐ None of the above |
| **Q4.** | (Narcolepsy only) People with type 1 narcolepsy experience cataplexy (a sudden loss of muscle tone triggered by strong emotions such as laughter, anger and surprise). People with type 2 narcolepsy don't. Do the patient notes specify the type of narcolepsy at diagnosis or whether cataplexy is experiences?<br>*(Please select one answer)*<br><br>☐ Type 1 narcolepsy or cataplexy<br>☐ Type 2 narcolepsy or no cataplexy<br>☐ No information available<br>☐ Other/unclear |

Once you have completed the two questions above, and are ready to submit the questionnaire, please select 'Complete' from the dropdown list below and then click to 'Save'.

Please note: Once you have marked the questionnaire as 'Complete' and saved you will no longer be able to edit your responses.

| **OBSTRUCTIVE SLEEP APNOEA SURVEY FORM** | |
|---|---|
| **Q1A.** | Has this patient been diagnosed or treated for the specified sleep disorder (see above) by a hospital specialist at any time?<br><br>☐ Yes *(Go to Q1B, Q1C and Q2A.)*<br>☐ No *(Go to Q3A.)* |
| **Q1B.** | When was the patient diagnosed by a hospital specialist?<br><br>☐ *(insert date DD/MM/YYYY – cannot choose a date in the future, the latest date should be current date)* |

| | |
|---|---|
| **Q1C.** | Which of the following methods were used to diagnose the patient?<br>*(Please select all that apply)*<br><br>☐ Electroencephalogram (EEG) telemetry<br>☐ Polysomnography<br>☐ Hospital respiratory polygraphy<br>☐ Multiple Sleep Latency Test (MSLT)<br>☐ Home respiratory polygraphy<br>☐ Home oximetry<br>☐ Successful Continuous positive airway pressure therapy (CPAP) trial<br>☐ Lumbar puncture<br>☐ Patient history<br>☐ Unclear or no information available<br>☐ None of the above |
| **Q2A.** | Has the diagnosis of the specified condition since been excluded by a hospital specialist and not been re-instated?<br><br>☐ Yes *(Go to Q5.)*<br>☐ No *(Go to Q5.)* |
| **Q3A.** | Has the patient been referred to a specialist for diagnosis?<br><br>☐ Yes *(Go to Q3B and Q3C.)*<br>☐ No *(Go to 3D.)* |
| **Q3B.** | When was the patient's most recent referral to a specialist for diagnosis?<br><br>☐ *(insert date DD/MM/YYYY – cannot choose a date in the future, the latest date should be current date)* |
| **Q3C.** | What was the result of the referral? *(Please select all that apply)*<br><br>☐ Patient on waiting list for initial appointment<br>☐ Patient undergoing investigation at hospital<br>☐ Diagnosis of specified condition excluded by hospital specialist<br>☐ Diagnosed with OSA (relevant when the specified sleep disorder is narcolepsy)<br>☐ Diagnosed with unspecified sleep apnoea<br>☐ Diagnosed with other sleep apnoea (Primary or central)<br>☐ Diagnosed with other sleep disorder<br>☐ Referral rejected |
| **Q3D.** | Which of the following information about the specified condition is included in the patient's record? (Please select all that apply)<br><br>☐ Suspected narcolepsy/OSA/sleep apnoea<br>☐ OSA/sleep apnoea/narcolepsy diagnosed by GP |

| | |
|---|---|
| | ☐ Patient request not to be referred for diagnosis<br>☐ GP decision not to refer for diagnosis<br>☐ None of the above |
| **Q5.** | (OSA only) What information, if any, is available about the severity of sleep apnoea at diagnosis? This may be in the form of an Apnoea Hypopnea Index (AHI) score or an oxygen desaturation index (ODI) score.<br>(Please select one answer)<br><br>☐ Mild (AHI 5 to 14)<br>☐ Moderate (AHI 15 to 30)<br>☐ Severe (AHI >30)<br>☐ Oxygen Desaturation Index score with severity not stated *(GO TO Q5A)*<br>☐ Unclear / no information available |
| **Q5A.** | Please enter the ODI score<br>[ ] *only accept numeric values)* |

Once you have completed the two questions above, and are ready to submit the questionnaire, please select 'Complete' from the dropdown list below and then click to 'Save'.

Please note: Once you have marked the questionnaire as 'Complete' and saved you will no longer be able to edit your responses.

### SA Table 1: Data sources, versions and DOIs

| **Database** | **Version** | **DOI** |
|---|---|---|
| CPRD Aurum | 2023.09.001 | https://doi.org/10.48329/6j2c-nh78 |
| CPRD Aurum HES APC | 2022.01.001 | https://doi.org/10.48329/vagx-9d96 |
| CPRD Aurum HES Outpatient | 2021.08.001 | https://doi.org/10.48329/7hm3-gt75HES |
| CPRD Aurum ONS deaths | 2022.01.001 | https://doi.org/10.48329/Q34F-F505 |

| | | |
|---|---|---|
| CPRD Aurum Small Area data (practice) – Carstairs Index | 2022.01.001 | https://doi.org/10.48329/3saw-rz63 |
| CPRD Aurum Small Area data (patient) – rural-urban classification | 2022.01.001 | https://doi.org/10.48329/aytt-h222 |

### SA Table 2: Variable definitions

| Variable | Type | Categories/units | Data source | Definition |
|---|---|---|---|---|
| **Recording of sleep disorder diagnosis** | | | | |
| Most specific code type on recorded diagnosis date | binary | OSA code, sleep apnoea code | CPRD Aurum/ HES APC | Most specific sleep apnoea code recorded on index date. OSA codes include codes for OSA and OSAS. SAS is included in OSA. Most codes are for OSA or SA. |
| Source of diagnostic code | categorical | CPRD, HES APC | CPRD Aurum/ HES APC | Data source in which sleep disorder is recorded on the recorded diagnosis date. CPRD Aurum = primary care, HES APC = inpatient hospital activitity data. There is an index code for both sources in a small number of records (<2%). These are coded as primary care to avoid small cell counts |
| Year of recorded diagnosis | continuous | calendar years (1998-2021) | CPRD Aurum/ HES APC | Recorded diagnosis date year |
| Year of recorded diagnosis compared to median in incident cohort | binary | before, after | CPRD Aurum/ HES APC | Before or after than the median calendar year in the incident cohort. Before category includes the median. |
| **Characteristics** | | | | |
| Age at recorded diagnosis | continuous | years | CPRD Aurum | age at index (estimated diagnosis date). Formular = 01/07/birthyear - indexdate |
| Age category | categorical | <9, < 18, 10-year categories, >85 | CPRD Aurum | see above. <9 and < 18 are defined for narcolepsy only. |
| Age compared to median in incident cohort | binary | younger, older | CPRD Aurum | younger or older than the median age in the incident cohort. Younger category includes the median. |
| Sex | binary | male, female | CPRD Aurum | sex recorded in GP practice |

| | | | | |
|---|---|---|---|---|
| Body Mass Index | categorical | Underweight/normal, overweight, obesity class I, obesity class II, obesity class III+, missing | CPRD Aurum | BMI values estimated from height, weight and BMI values in observation file (see algorithm in Github site for details) in observation. Categorised using World Health Organisation categories. Underweight and healthy normal weight combined to avoid small cell counts. Used nearest value to index date. Missing if BMI not measured on or prior to index. |
| Body Mass Index binary | binary | obesity class I or under, obesity class II+ | CPRD Aurum | as above |
| Ethnicity | categorical | White, South Asian, Black, Other, Mixed | CPRD Aurum/ HES (all) | Derived using the most commonly recorded ethnicity in primary care data (or latest if equally common). Unknown and missing values replaced with most commonly reported ethnicity in HES where available. See Github site for more detail. |
| Practice area-based deprivation quintiles | categorical | 1 (least deprived), 2, 3, 4, 5 (most deprived) | Carstairs Index linked to the 2011 census | linked to practice postcode. Quintiles are estimated by CPRD using national Carstairs data. |
| Practice area-based deprivation binary | binary | less deprived, more deprived | Carstairs Index linked to the 2011 census | less deprived= quintiles 1 to 3, most deprived = quintiles 4 and 5 |
| Patient urban-rural status | binary | urban, rural | Rural Urban Classifications (RUC) for England and Wales for 2011 census population | linked to patient postcode. Missing if practice not linked to rural/urban data in census. |
| Practice size | continuous | registered patients | CPRD Aurum | Practice size for each practice estimated using full study population for incidence/prevalence study in mid-2019 or the year prior to the index date for practices leaving CPRD prior to mid-2019. |
| Practice size relative to median in incident cohort | binary | smaller, larger | CPRD Aurum | smaller or larger than the median practice size in the incident cohort. The smaller category includes the median |

| | | | | |
|---|---|---|---|---|
| **Additional information in routinely collected data** | | | | |
| Outpatient visits (any) | Binary | Yes/No | HES Outpatient data | Attended at least one outpatient visit in the 6 months before or after index |
| Neurology outpatient attendance | Binary | Yes/No | HES Outpatient data | Attended at least one neurology consultant outpatient visit in the 6 months before or after index (includes neurology NOS, paediatric neurology & clinical neurophysiology) |
| Respiratory outpatient attendance | Binary | Yes/No | HES Outpatient data | Attended at least one respiratory consultant outpatient visit in the 6 months before or after index |
| Paediatric outpatient attendance | Binary | Yes/No | HES Outpatient data | Attended at least one paediatric (NOS) consultant outpatient visit in the 6 months before or after index |
| Ear, Nose and Throat (ENT) outpatient attendance | Binary | Yes/No | HES Outpatient data | Attended at least one ENT consultant outpatient visit in the 6 months before or after index |
| Anaesthetist outpatient attendance | Binary | Yes/No | HES Outpatient data | Attended at least one anaesthetic consultant outpatient visit in the 6 months before or after index |
| Sleep-related consultants combined | Binary | Yes/No | HES Outpatient data | Attended at least one outpatient visit to possible sleep medicine consultant in the 6 months before or after index (i.e. neurology, respiratory, paediatric, ENT, or anaesthetist) |
| EDS drug prescription | Binary | Yes/No | CPRD Aurum data | At least one prescription of an EDS drug at any time in patient record. Drugs included modafinil, methylphenidate, and dexamfetamine (excluded lisdexamfetamine). Code lists were developed using lower case search to search drug substance, product name, and termfromemis fields - terms recorded anywhere in string. Inclusion terms: modaf provigil methylphenidate concerta delmosart equasym medikinet xaggitin tranquilyn ritalin affenid matoride xenidate equasym meflynate metyrol dexam, amfexa, Dexedrine) Exclusion terms (lisdex adderall durophet). See Github for more detail. |
| **Validation questionnaire (transformed variables)** | | | | |

| | | | | |
|---|---|---|---|---|
| Recorded role of hospital specialist (summary) | Categorical | Diagnosis or treatment of sleep disorder, Referral but no diagnosis, No referral | Validation questionnaire responses | Diagnosis or treatment of sleep disorder (Q1a=1), Referral but no diagnosis (Q3a = 1), No referral (Q3a=0) |
| Days between recorded and specialist diagnosis date (positive = recorded later) | continuous | days | CPRD Aurum, HES APC, Validation questionnaire responses | Recorded diagnosis date - q1b (Date of diagnosis by hospital specialist) |
| Months between the recorded and specialist diagnosis date | categorical | >6 months before, 1 to 6 months before, within 1 month, 1 to 6 months after, > 6 months after | CPRD Aurum, HES APC, Validation questionnaire responses | Difference between recorded and diagnosis date categorised (month = 30 days, 6 months = 183 days) |
| Diagnosis recorded within +/- 6 months of specialist diagnosis date | binary | More than 6 months, within 6 months | CPRD Aurum, HES APC, Validation questionnaire responses | Difference between the recorded and diagnosis date as a binary variable |
| Months between recorded diagnosis date and referral date (positive = index later) | continuous | days | CPRD Aurum, HES APC, Validation questionnaire responses | (Recorded diagnosis date - q3b (Date of referral to hospital specialist))/30 |
| OSA severity | Categorical | Mild (AHI 5 to 14), Moderate (AHI 15 to 30), Severe (AHI >30), Unclear / no information available | Validation questionnaire responses | Combined Q4 (severity measured using AHI scores) and Q5a (ODI score). ODI scores classified using the same thresholds as ADI. https://www.ncbi.nlm.nih.gov/pmc/articles/PMC8889990/ |

| | | | | |
|---|---|---|---|---|
| At least one objective diagnostic method identified | Binary | Yes/No | Validation questionnaire responses | At least one of the following tests recorded: OSA and narcolepsy (q1c1 Electroencephalogram (EEG) telemetry, q1c2 Polysomnography, q1c4 Multiple Sleep Latency Test (MSLT)), OSA only (q1c3 Hospital respiratory polygraphy, q1c5 Home respiratory polygraphy, q1c6 Home oximetry, q1c7 Successful Continuous positive airway pressure therapy (CPAP) trial), narcolepsy only (q1c8 lumbar puncture) |

### SA Table 3: Diagnostic and demographic characteristics of gold standard and false cases

| | OSA false | OSA gold standard | narcolepsy false | narcolepsy gold standard |
|---|---|---|---|---|
| **N+** | 40 | 104 | 42 | 59 |
| **RECORDING OF SLEEP DISORDER DIAGNOSIS** | | | | |
| **Source of diagnostic code*** | | | | |
| CPRD Aurum | 15 (37.5) | 80 (76.9) | 20 (47.6) | 42 (71.2) |
| HES APC | 25 (62.5) | 24 (23.1) | 22 (52.4) | 17 (28.8) |
| **Most specific code type recorded on diagnosis date** | | | | |
| OSA code | >35 (>87.5) | 58 (55.8) | | |
| sleep apnoea | <5 (<12.5) | 46 (44.2) | | |
| missing | 0 (0.0) | 0 (0.0) | | |
| **Person-years before diagnosis date** | | | | |
| Mean (SD) | 17.4 (12.6) | 16.1 (14.0) | 13.4 (11.8) | 14.2 (11.3) |
| Median (IQR) | 16.3 (5.6, 23.0) | 12.9 (4.7, 23.6) | 10.7 (4.9, 19.0) | 12.8 (5.4, 20.6) |
| **Year of recorded diagnosis** | | | | |
| Mean (SD) | 2013.4 (6.4) | 2014.3 (4.8) | 2013.8 (6.4) | 2012.3 (6.6) |
| Median (IQR) | 2015 (2009, 2019) | 2016 (2012, 2018) | 2015.0 (2011, 2019) | 2014.0 (2008, 2018) |
| **ADDITIONAL INFORMATION IN ROUTINELY COLLECTED DATA** | | | | |
| **Outpatient visit within 6 months of recorded diagnosis (HES OP data)** | | | | |
| Linked OP data available | 32 (80.0) | 97 (93.3) | 32 (76.2) | 43 (72.9) |
| All | 24 (75.0) | 86 (88.7) | 27 (84.4) | 37 (86.0) |
| Neurology | <5 (<15.6) | <5 (<5.2) | 13 (40.6) | 22 (51.2) |
| Respiratory | 6 (18.8) | 50 (51.5) | 10 (31.2) | 13 (30.2) |
| Paediatric (NOS) | 0 (0.0) | 0 (0.0) | <5 (<15.6) | 5 (11.6) |

| | | | | |
|---|---|---|---|---|
| Ear Nose & Throat | 5 (15.6) | 18 (18.6) | <5 (<15.6) | <5 (<11.6) |
| Anaesthetics | 0 (0.0) | 0 (0.0) | <5 (<15.6) | 5 (11.6) |
| Sleep-related consultants combined | 10 (31.2) | 59 (60.8) | 21 (65.6) | 29 (67.4) |
| EDS drug prescription ever | | | 7 (16.7) | 42 (71.2) |
| **CHARACTERISTICS** | | | | |
| **Age at recorded diagnosis (years)** | | | | |
| Mean (SD) | 54.6 (12.4) | 51.7 (12.3) | 38.9 (15.6) | 36.7 (18.3) |
| Median (IQR) | 56.2 (46.7, 64.0) | 53.0 (44.0, 61.0) | 41.0 (30.0, 48.9) | 39.0 (20.7, 48.8) |
| **Sex** | | | | |
| male | 28 (70.0) | 76 (73.1) | 20 (47.6) | 28 (47.5) |
| female | 12 (30.0) | 28 (26.9) | 22 (52.4) | 31 (52.5) |
| **Body Mass Index** | | | | |
| Under/normal weight | 5 (12.5) | 8 (7.7) | | |
| Overweight | 11 (27.5) | 16 (15.4) | | |
| Obesity class I | 11 (27.5) | 31 (29.8) | | |
| Obesity class II | <5 (<12.5) | 25 (24.0) | | |
| Obesity class III+ | 6 (15.0) | 23 (22.1) | | |
| missing | <5 (<12.5) | <5 (<4.8) | | |
| **Ethnicity** | | | | |
| White | 35 (87.5) | 91 (87.5) | 35 (83.3) | 53 (89.8) |
| South Asian | <5 (<12.5) | 5 (4.8) | <5 (<11.9) | <5 (<8.5) |
| Black | <5 (<12.5) | 8 (7.7) | <5 (<11.9) | <5 (<8.5) |
| Other | 0 (0.0) | 0 (0.0) | <5 (<11.9) | <5 (<8.5) |
| Mixed | <5 (<12.5) | 0 (0.0) | 0 (0.0) | 0 (0.0) |
| missing | 0 (0.0) | 0 (0.0) | <5 (<11.9) | 0 (0.0) |
| **Carstairs quintile** | | | | |
| 1 (least deprived) | 8 (20.0) | 24 (23.1) | 10 (23.8) | 14 (23.7) |
| 2 | 9 (22.5) | 18 (17.3) | 5 (11.9) | 13 (22.0) |
| 3 | 9 (22.5) | 25 (24.0) | 13 (31.0) | 13 (22.0) |

| | | | | |
|---|---|---|---|---|
| 4 | 7 (17.5) | 18 (17.3) | 8 (19.0) | 10 (16.9) |
| 5 (most deprived) | 7 (17.5) | 19 (18.3) | 6 (14.3) | 9 (15.3) |
| **Urban Rural** | | | | |
| urban | 30 (75.0) | 83 (79.8) | 31 (73.8) | 47 (79.7) |
| rural | 10 (25.0) | 21 (20.2) | 11 (26.2) | 12 (20.3) |
| **Practice size** | | | | |
| Mean (SD) | 39012.4 (27532.8) | 42149.0 (35771.9) | 39485.8 (31987.1) | 46252.7 (40352.5) |
| Median (IQR) | 30926.0 (20011.5, 47620.5) | 31269.0 (21171.0, 45511.0) | 30706.0 (17338.0, 44124.0) | 33259.0 (24635.0, 52162.0) |

### SA Table 4. Crude PPV stratified by characteristics

| Covariate | Covariate value | Gold standard cases (n) | Sample (n) | PPV (95% CI) | PPV ratio (95% CI) | PPV ratio p-value |
|---|---|---|---|---|---|---|
| **OSA (primary algorithm)** | | | | | | |
| Sex | male | 76 | 104 | 73.1 (63.5-81.3) | 1 | |
| | female | 28 | 40 | 70.0 (53.5-83.4) | 0.96 (0.76-1.21) | 0.72 |
| Age compared to median in incident cohort | Younger | 50 | 65 | 76.9 (64.8-86.5) | 1 | |
| | older | 54 | 79 | 68.4 (56.9-78.4) | 0.89 (0.73-1.09) | 0.25 |
| Practice area-based deprivation | Less deprived | 67 | 93 | 72.0 (61.8-80.9) | 1 | |
| | More deprived | 37 | 51 | 72.5 (58.3-84.1) | 1.01 (0.81-1.24) | 0.95 |
| BMI binary | Obesity Class I and below | 55 | 82 | 67.1 (55.8-77.1) | 1 | |
| | Obesity class II plus | 48 | 58 | 82.8 (70.6-91.4) | 1.23 (1.02-1.50) | 0.03 |
| Year of recorded diagnosis compared to median in incident cohort | Before | 57 | 79 | 72.2 (60.9-81.7) | 1 | |
| | After | 47 | 65 | 72.3 (59.8-82.7) | 1.00 (0.82-1.23) | 0.98 |
| Practice size compared to median in incident cohort | Smaller | 56 | 78 | 71.8 (60.5-81.4) | 1 | |
| | larger | 48 | 66 | 72.7 (60.4-83.0) | 1.01 (0.83-1.24) | 0.90 |
| **Narcolepsy (primary algorithm)** | | | | | | |
| Sex | male | 28 | 48 | 58.3 (43.2-72.4) | 1 | |
| | female | 31 | 53 | 58.5 (44.1-71.9) | 1.00 (0.72-1.40) | 0.99 |
| Age compared to median in incident cohort | Younger | 27 | 45 | 60.0 (44.3-74.3) | 1 | |
| | older | 32 | 56 | 57.1 (43.2-70.3) | 0.95 (0.68-1.33) | 0.77 |
| Practice area-based deprivation | Less deprived | 40 | 68 | 58.8 (46.2-70.6) | 1 | |
| | More deprived | 19 | 33 | 57.6 (39.2-74.5) | 0.98 (0.69-1.40) | 0.91 |
| Year of recorded diagnosis compared to median in incident cohort | Before | 34 | 51 | 66.7 (52.1-79.2) | 1 | |
| | After | 25 | 50 | 50.0 (35.5-64.5) | 0.75 (0.53-1.05) | 0.10 |
| Practice size compared to median in incident cohort | Smaller | 28 | 54 | 51.9 (37.8-65.7) | 1 | |
| | larger | 31 | 47 | 66.0 (50.7-79.1) | 1.27 (0.91-1.77) | 0.15 |

### SA Table 5: Recorded role of hospital specialist and description of gold standard cases, by algorithm

| **OSA** | **Primary (CPRD or HES APC)** | **CPRD** | **CPRD & HES APC** | **Primary with OP visit to sleep-related specialty** | **CPRD with OP visit to sleep-related specialty** |
|---|---|---|---|---|---|
| **N+** | **144** | **109** | **51** | **69** | **55** |
| **ALL CASES** | | | | | |
| **Recorded role of hospital specialist** | | | | | |
| Diagnosis or treatment of sleep disorder | 104 (72.2) | 93 (85.3) | 48 (94.1) | 59 (85.5) | 54 (98.2) |
| Referral but no diagnosis | 14 (9.7) | 6 (5.5) | 0 (0.0) | <5 (<7.2) | 0 (0.0) |
| No referral | 26 (18.1) | 10 (9.2) | <5 (<9.8) | 7 (10.1) | <5 (<9.1) |
| **CASES DIAGNOSED OR TREATED BY A HOSPITAL SPECIALIST** | | | | | |
| **Days between the recorded and specialist diagnosis date (positive = recorded later)** | | | | | |
| Mean (SD) | -137.7 (654.4) | -152.7 (667.5) | 469.2 (820.7) | -91.5 (450.6) | -75.4 (508.8) |
| Median (IQR) | -1.0 (-72.0, 0.0) | 0.0 (-62.0, 0.0) | 220.5 (1.0, 695.5) | 0.0 (-45.0, 16.0) | 0.0 (-45.0, 11.0) |
| **Recorded data within 6 months before of after specialist date** | 83 (80.6) | 75 (81.5) | 22 (45.8) | 53 (89.8) | 47 (87.0) |
| **At least one objective diagnostic method identified** | 81 (77.9) | 74 (79.6) | 39 (81.2) | 45 (76.3) | 42 (77.8) |
| **Severity of OSA (AHI or ODI)** | | | | | |
| Mild (AHI 5 to 14) | 22 (21.2) | 19 (20.4) | 11 (22.9) | 15 (25.4) | 14 (25.9) |
| Moderate (AHI 15 to 30) | 29 (27.9) | 28 (30.1) | 15 (31.2) | 14 (23.7) | 14 (25.9) |
| Severe (AHI >30) | 32 (30.8) | 30 (32.3) | 17 (35.4) | 19 (32.2) | 17 (31.5) |
| Unclear / no information available | 21 (20.2) | 16 (17.2) | 5 (10.4) | 11 (18.6) | 9 (16.7) |
| **Diagnosis excluded by specialist at a later date** | 10 (9.6) | 8 (8.6) | <5 (<10.4) | 5 (8.5) | 5 (9.3) |

| Narcolepsy | Primary (CPRD or HES APC) | CPRD | CPRD & HES APC | Primary with OP visit to sleep-related specialty | CPRD with OP visit to sleep-related specialty | CPRD with EDS drug prescription | CPRD with EDS drug prescription |
|---|---|---|---|---|---|---|---|
| **N+** | **101** | **69** | **28** | **50** | **34** | **49** | **45** |
| **ALL CASES** | | | | | | | |
| **Recorded role of hospital specialist** | | | | | | | |
| Diagnosis or treatment of sleep disorder | 59 (58.4) | 49 (71.0) | 26 (92.9) | 29 (58.0) | 27 (79.4) | 42 (85.7) | 40 (88.9) |
| Referral but no diagnosis | 22 (21.8) | 7 (10.1) | <5 (<17.9) | 14 (28.0) | <5 (<14.7) | <5 (<10.2) | <5 (<11.1) |
| No referral | 20 (19.8) | 13 (18.8) | 0 (0.0) | 7 (14.0) | <5 (<14.7) | <5 (<10.2) | <5 (<11.1) |
| **CASES DIAGNOSED OR TREATED BY A HOSPITAL SPECIALIST** | | | | | | | |
| **Days between the recorded and confirmed diagnosis date (positive = recorded later)** | | | | | | | |
| Mean (SD) | 503.6 (2526.5) | 154.0 (1152.3) | 1809.3 (2183.6) | -168.8 (1152.3) | -81.3 (1277.9) | 352.9 (1338.0) | 280.6 (1306.4) |
| Median (IQR) | 0.0 (-71.0, 273.0) | 0.0 (0.0, 143.0) | 1232.5 (0.0, 3085.0) | 0.0 (-102.0, 21.0) | 0.0 (-17.0, 88.0) | 88.0 (21.0, 978.0) | 79.0 (14.5, 616.5) |
| **Recorded data within 6 months before of after confirmed date** | 37 (62.7) | 33 (67.3) | 8 (30.8) | 19 (65.5) | 17 (63.0) | 20 (47.6) | 20 (50.0) |
| **At least one objective diagnostic method identified** | 35 (59.3) | 30 (61.2) | 17 (65.4) | 19 (65.5) | 19 (70.4) | 28 (66.7) | 26 (65.0) |
| **Type of narcolepsy** | | | | | | | |
| Type 1 narcolepsy or cataplexy | 18 (30.5) | 16 (32.7) | 9 (34.6) | 10 (34.5) | 10 (37.0) | 14 (33.3) | 14 (35.0) |
| Type 2 narcolepsy or no cataplexy | 12 (20.3) | 11 (22.4) | 6 (23.1) | 6 (20.7) | 6 (22.2) | 11 (26.2) | 10 (25.0) |
| No information available | 16 (27.1) | 11 (22.4) | 5 (19.2) | 6 (20.7) | 5 (18.5) | 6 (14.3) | 6 (15.0) |
| Other/unclear | 13 (22.0) | 11 (22.4) | 6 (23.1) | 7 (24.1) | 6 (22.2) | 11 (26.2) | 10 (25.0) |
| **Diagnosis excluded by specialist at a later date** | 5 (8.5) | <5 (<10.2) | <5 (<19.2) | <5 (<17.2) | <5 (<18.5) | <5 (<11.9) | <5 (<12.5) |

### SA Figure 1: Incidence rate by months since registration in CPRD practice

Caption: OSA = Obstructive Sleep Apnoea. Black dotted line = 3 months since registration

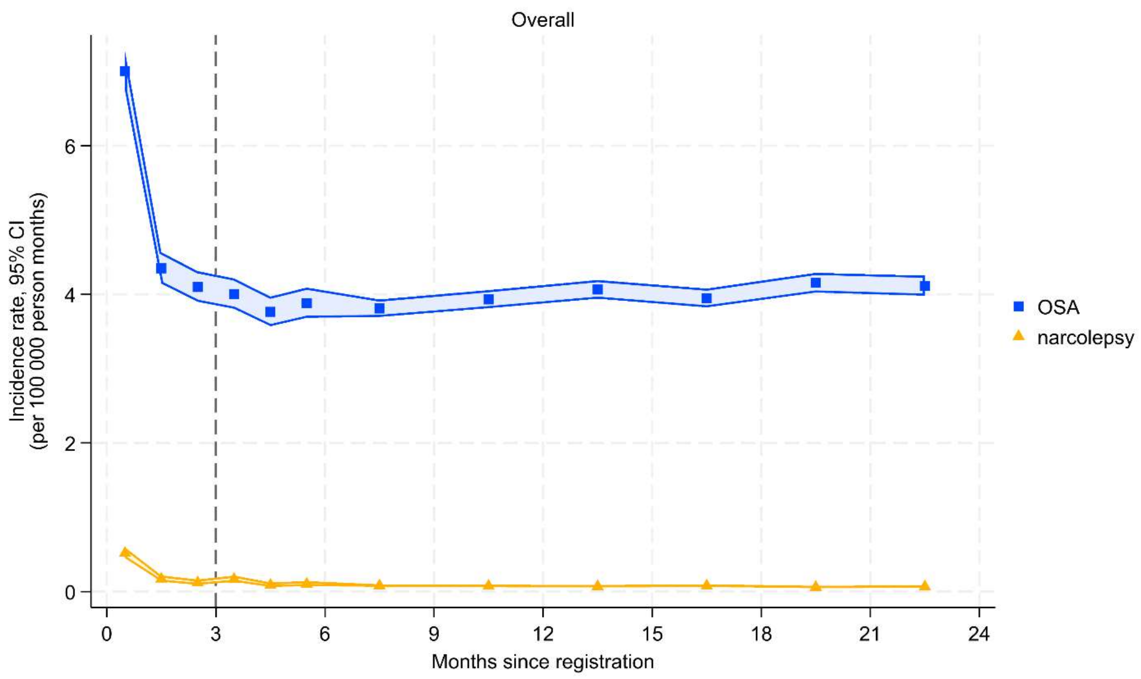

### SA Figure 2: Participant flow diagram

Note: To restrict the study population to people contributing data in September 2023, we excluded practices who last contributed data before May 2023. This allows for time taken to process the data between data collection and database release.

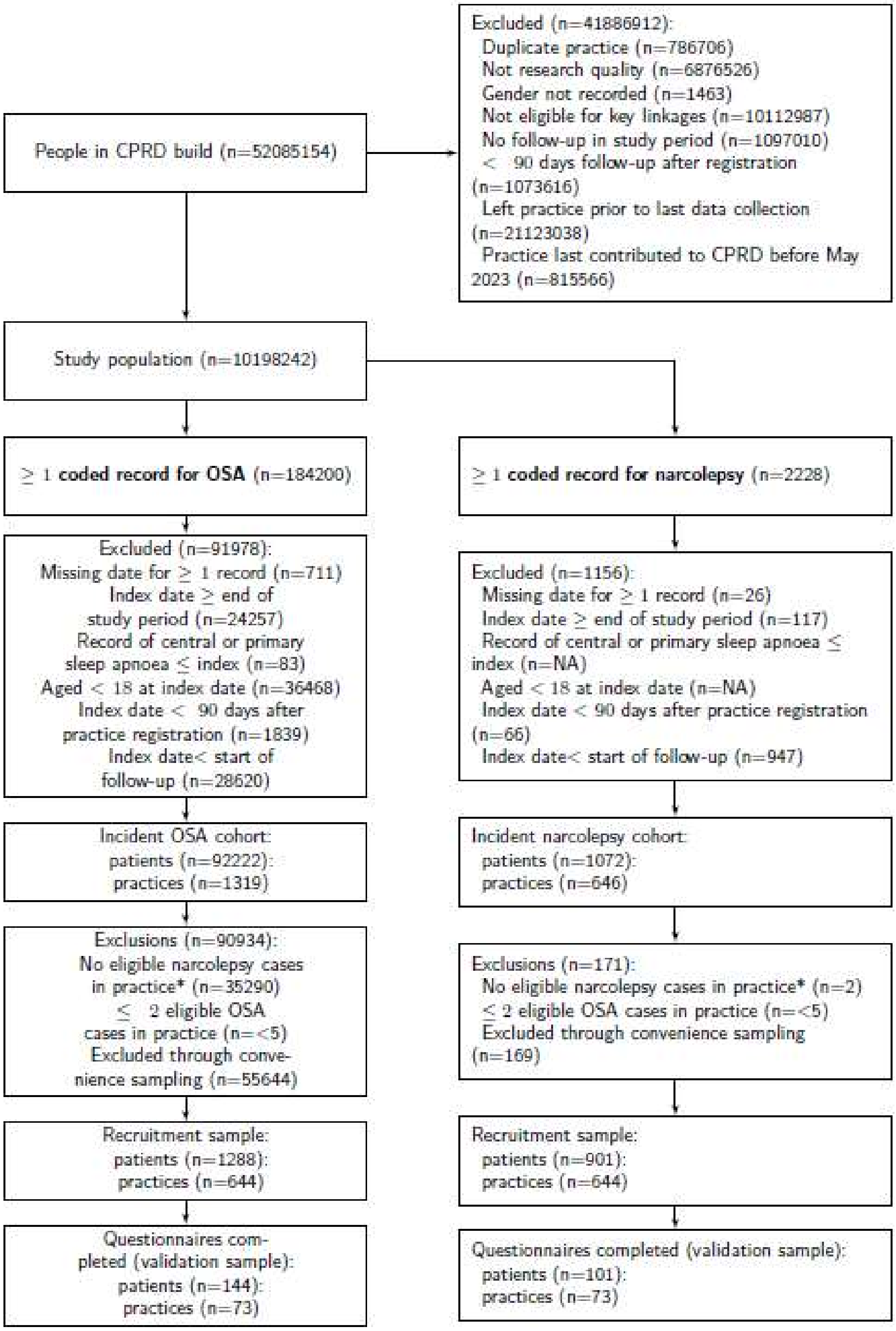
